# Using predicted imports of 2019-nCoV cases to determine locations that may not be identifying all imported cases

**DOI:** 10.1101/2020.02.04.20020495

**Authors:** PM De Salazar, R Niehus, A Taylor, C Buckee, M Lipsitch

**Author notes:** **Correspondence:** Pablo M. De Salazar. Center for Communicable Disease Dynamics, Department of Epidemiology, Harvard T.H. Chan School of Public Health,677 Huntington Ave Boston, Massachusetts, United States of America. Marc Lipsitch. Center for Communicable Disease Dynamics, Department of Epidemiology, Harvard T.H. Chan School of Public Health, 677 Huntington Ave Boston, Massachusetts, United States of America. contributed equally (PM De Salazar, R Niehus).

## Abstract

Cases from the ongoing outbreak of atypical pneumonia caused by the 2019 novel coronavirus (2019-nCoV) exported from mainland China can lead to self-sustained outbreaks in other populations. Internationally imported cases are currently being reported in several different locations. Early detection of imported cases is critical for containment of the virus. Based on air travel volume estimates from Wuhan to international destinations and using a generalized linear regression model we identify locations which may potentially have undetected internationally imported cases.

## Introduction

A novel coronavirus 2019-nCoV has been identified with the first confirmed patient cases in December 2019 in the city of Wuhan, capital of Hubei province, China. Since then the number of confirmed cases has increased drastically. Model estimates suggested that by Jan 25, 2020 there were a total of over 75,000 infected cases with a doubling time of about 6 days^1^. By the end of January 2020, city-wide lockdowns were implemented for Wuhan and neighbouring cities. Nonetheless, the virus has managed to spread from Wuhan to other Chinese cities and also outside of China. As of Feb 4, 2020, cases outside China were being reported in 23 different locations of which 22 locations had imported cases (Spain reported one case but due to secondary transmission)^2^.

The majority of these cases have been linked to a recent travel history from China^2^, suggesting that air travel may play an important role for the risk of cases being exported outside of China. To prevent other cities and countries becoming epicentres of the 2019-nCoV epidemic, substantial targeted public health interventions are required, first for detection of cases and then for control of local spread. Here we use estimates of air travel volume from Wuhan to international destinations in 49 locations expected to be proficient at detecting imported cases based on having a high Global Health Security (GHS) Index^3^, and a generalised linear regression model to predict imports of 2019-nCoV cases across 191 locations. Using these predictions we can identify locations that might not be identifying imported cases.

## Methods

Under a model fit to data from destinations with high surveillance capacity, we sought to identify locations that report fewer than predicted imported cases of 2019-nCoV. Specifically, for 49 locations with high surveillance capacity we regressed the cumulative number of confirmed imported cases of 2019-nCoV in international destinations outside mainland China on the estimated number of daily air flight passengers coming from Wuhan airport by estimated direct or indirect international flights (daily air travel volume) using a Poisson model. We then use predictions from this model to compare to reported cases across all 191 locations.

The model requires three types of data: data on imported cases of 2019-nCoV, data on daily air travel volume, and data on surveillance capacity. Data on imported cases aggregated by destination were obtained from the WHO technical report dated 4th February 2020^2^ (a zero case count was assumed for all locations not listed). We used case counts up to the 4th February, because after this date the number of exported cases from Hubei drops rapidly^2^, likely due to the Hubei-wide lockdowns. We defined imported cases as those with known travel history from China (of those, 83% had travel history from Hubei province, and 17% from unknown locations in China^2^). We excluded cases that are likely due to transmission outside of China or whose transmission source is still under investigation^2^. In addition, we excluded from our model Hong Kong, Macau and Taiwan because locally transmitted and imported cases were not disaggregated in these locations. In summary, of 195 worldwide locations (in general, locations reflect countries without any position on territorial claims) our model considers N=191 due to the exclusion of China, Hong Kong, Macau and Taiwan. Data on daily air travel volume was obtained as follows. Lai et al.^4^ report monthly air travel volume estimates for the 27 locations most connected to Wuhan outside mainland China. These estimates are based on historical (February 2018) data from the International Air Travel Association^4^. They include direct and indirect flight itineraries from Wuhan to destinations outside of China. For all 164 locations not listed by Lai et al.^4^, we set the daily air travel volume to 1.5 passengers per day, which is one half of the minimum reported by Lai et al. Surveillance capacity was assessed using the Global Health Security Index, which is an assessment of health security including detection and reporting capabilities across 195 countries agreeing to the International Health Regulations (IHR [2005])^3^. Specifically, we use the Early Detection and Reporting Epidemics of Potential International Concern component of the Index^3^, henceforth referred to as simply the GHS index, and define high surveillance locations as those whose GHS index is greater than the 75th quantile.

The model is as follows. We assumed that across the *n*=49 high surveillance locations the case counts follow a Poisson distribution, and that the expected case count is linearly proportional to the daily air travel volume:

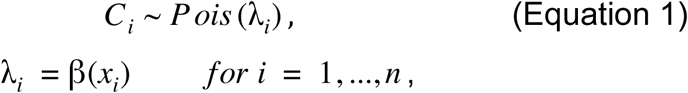

where *Ci* denotes the reported case count in the *i*-th location, *λi* denotes the expected case count in the *i*-th location, *β* denotes the regression coefficient, *xi* denotes the daily air travel volume of the *i*-th location.

The model was fitted in R(version 3.6.1)^5^ to compute 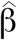, the maximum likelihood estimate of *β*, and thus the expected case count given high surveillance (Figure 1, solid grey line). We also computed the 95% prediction interval (PI) bounds under this model of high surveillance (Figure 1, dashed lines) for all *N*=191 values of daily air travel volume as follows. First, we generated a bootstrapped dataset by sampling *n* locations with replacement among high surveillance locations. Second, we re-estimated *β* using the bootstrapped dataset. Third we simulated case counts for all *N*=191 locations under our model using the estimate of *β* based on the bootstrapped dataset. These three steps were repeated 50,000 times to generate for each of the *N* locations 50,000 simulated case counts from which the lower and upper PI bounds (2.5th and 97.5th percentiles) were computed. In Figure 1 the 95% PI bounds were smoothed using functions from the R package ggplot2^6^. The reported case counts of all *n*=49 high surveillance locations (22 with non-zero case counts plus 27 with zero case counts) used to fit the model (Figure 1, purple-coloured points) are plotted alongside those for all 142 locations that do not have high surveillance (Figure 1, blue-coloured points).

**Figure 1.**
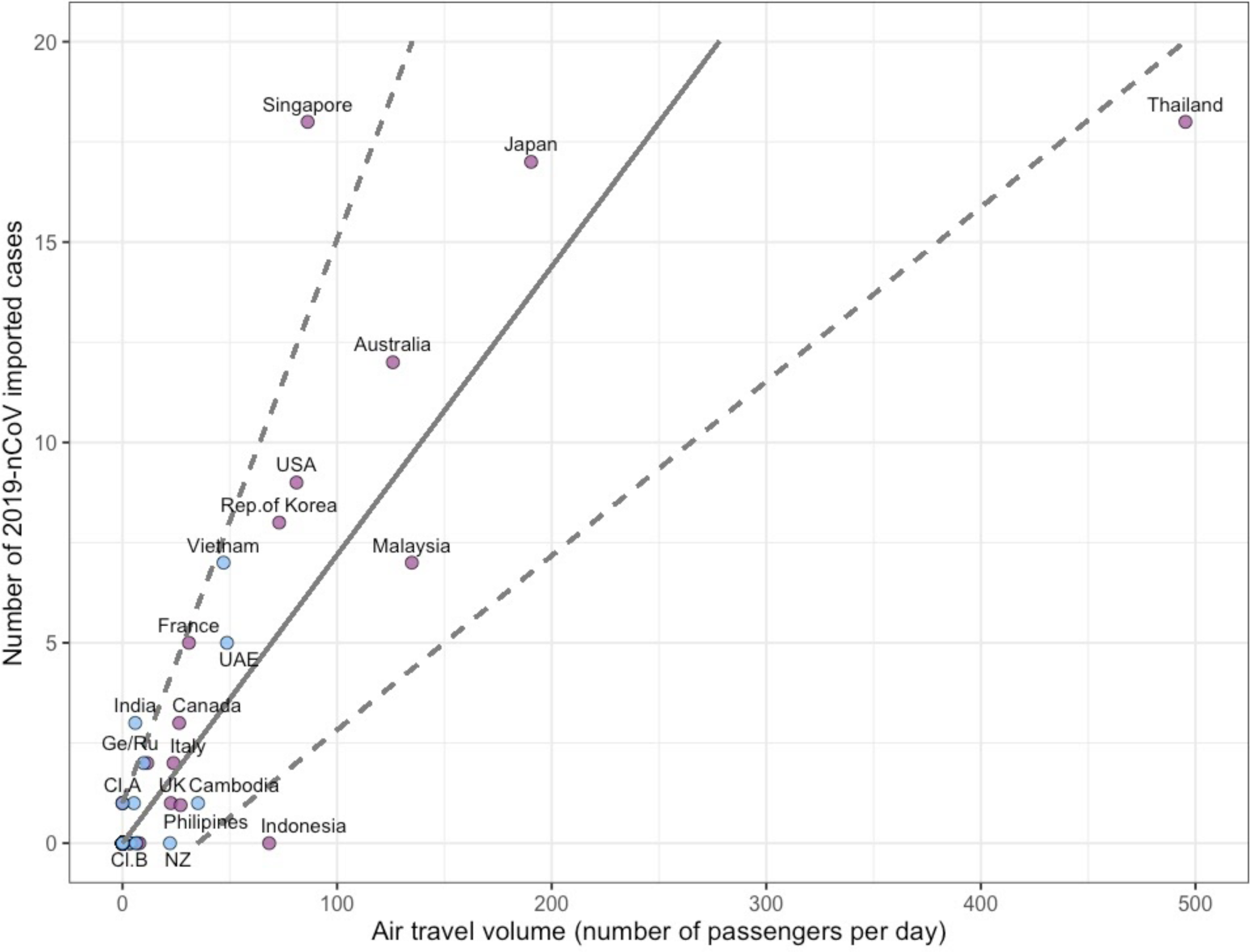
Imported case counts and daily air travel volume. The solid line shows the expected case counts based on our model fitted to high surveillance locations (purple points), and the dashed lines show the 95% prediction interval bounds for all locations including those that do not have high surveillance (light blue points). Cluster A (Cl.A) comprises locations with one imported case reported and air travel <10 passengers per day: Nepal, Sri Lanka, Finland, and Sweden. Cluster B (Cl.B) comprises locations (*n*=161) with no reported cases and estimated air travel <10 passengers per day. Other abbreviations: Germany (Ge), New Zealand (NZ), United Arab Emirates (UAE), United States of America (USA), United Kingdom (UK), Russia (Ru).

To assess the robustness of our results we reran the analysis as described above, but each time implementing one of the following changes: 1) setting the daily air travel volume for locations not listed by Lai et al. to 0.1, 1, or 3 (instead of 1.5) passengers per day; 2) defining high surveillance locations using a more lenient and a more stringent GHS index (50th and 95th quantile respectively instead of 75th); 3) excluding Thailand from the model fit since it is a high-leverage point. In total, we thus did six additional regression analyses (see Supplementary Figure 1). All analyses are fully reproducible given code available online (https://github.com/c2-d2/cov19flightimport).

## Results

Imported case counts of 2019-nCoV among high surveillance locations are positively correlated with daily air travel volume (Figure 1). An increase in flight volume by 14 passengers per day is associated with one additional imported case in expectation. Singapore lies above the 95% prediction interval (PI), with 12 (2-13) more reported import cases than expected under our model. Thailand has a relatively high air travel volume as compared to all other locations, yet it lies below the 95% prediction interval. Finally, Indonesia with zero reported cases lies below the prediction interval – its expected case count is 5 (1-11 95% PI). This overall pattern (Singapore lying above the 95% PI, Thailand and Indonesia below) was observed consistently across all six robustness regression analyses.

**Supplementary Figure 1.**
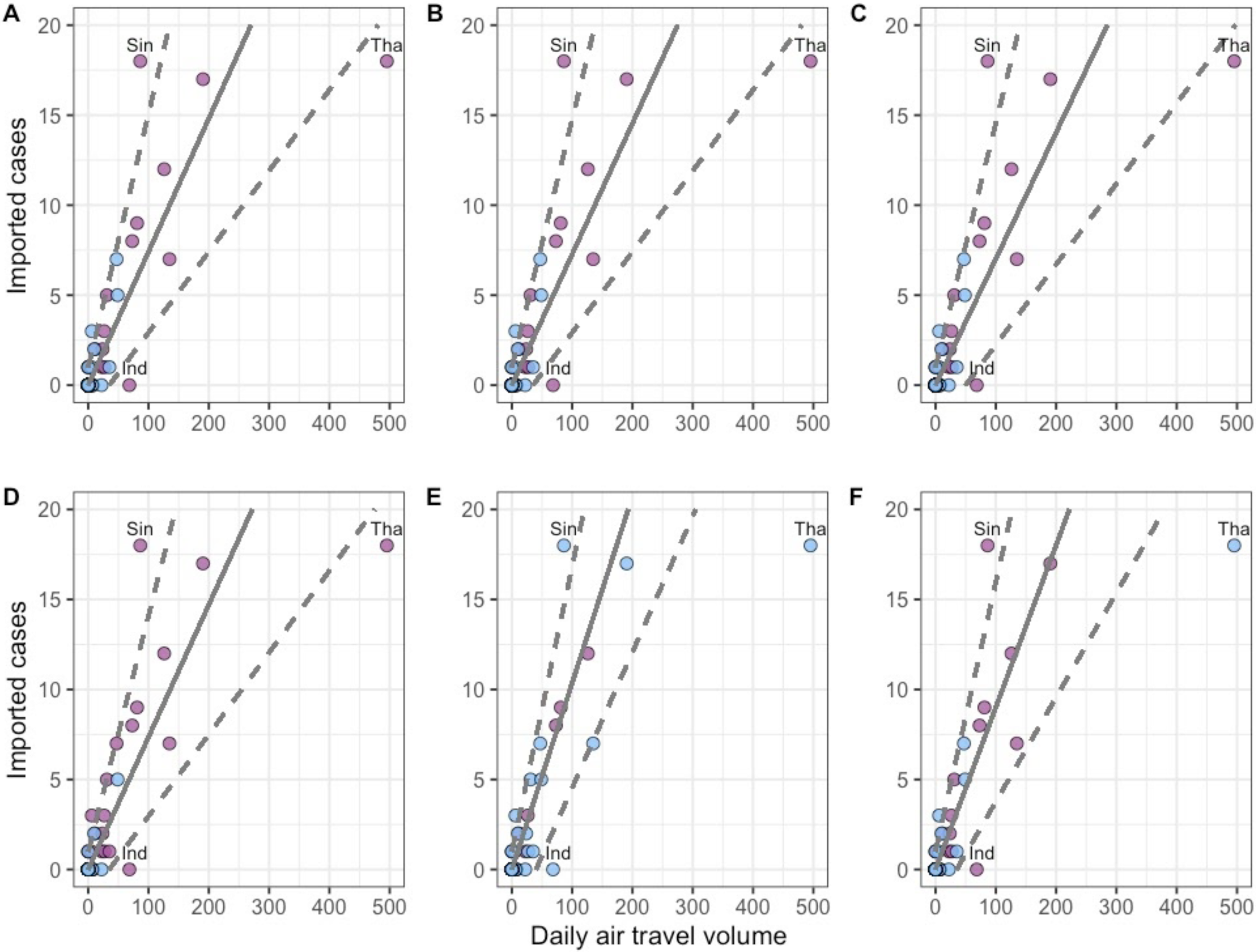
Imported case counts and daily air travel volume under six different regression analyses. Six different analyses are shown: **A**-**C**) setting the daily air travel volume for locations not listed by Lai et al. to 0.1, 1, or 3 passengers per day, respectively; **D**) defining high surveillance locations using a more lenient GHS index (50th quantile) to define high surveillance locations and **E**) a more stringent GHS index (95th quantile) to define high surveillance locations; **F**) excluding Thailand from the model fit. Throughout, the solid line shows the expected case counts based on our model fitted to high surveillance locations (purple points), and the dashed lines show the 95% prediction interval bounds for all locations including those without high surveillance (light blue points). Across all six regression analyses, Singapore lies above the 95% PI, Thailand and Indonesia below. Abbreviations: Indonesia (Ind), Thailand (Tha), Singapore (Sin).

## Discussion

We aimed to identify locations with underdetected cases by fitting a model to the cumulative international imported case counts of nCoV-2019 reported by high surveillance locations and Wuhan-to-location air travel volume. Our model can be adjusted to account for exportation of cases from locations other than Wuhan as the outbreak develops and more information on importations and self-sustained transmission becomes available. One key advantage of this model is that it does not rely on estimates of incidence or prevalence in the epicentre of the outbreak.

Based on our model, locations whose case counts exceed the 95% prediction interval (PI) could be interpreted as having higher case-detection capacity and/or more connection with Wuhan than that captured by available daily air travel volume, such as land transportation. Locations below the 95% PI may have undetected cases based on the expected case count under high surveillance. We recommend that outbreak surveillance and control efforts for potential local transmission should be rapidly strengthened in those locations lying below the 95% PI lower bound, in particular Indonesia, to ensure detection of cases and appropriate control measures to reduce the risk of self-sustained transmission.

## Data Availability

Code and data are available at https://github.com/c2-d2/cov19flightimport

